# Ready-To-Use Investigational Stem Cells, MiSaver, in Patients with Recent Acute Myocardial Infarction, 1 year follow up from a Phase 1 Safety Study

**DOI:** 10.1101/2024.04.01.24305125

**Authors:** KC Ueng, CF Tsai, CH Su, YT Chuang, JTK Liu

## Abstract

**Objectives:** The aim of this study was to investigate the safety of MiSaver stem cells and their preliminary efficacy in improving left ventricular ejection function and functional activity in patients with acute myocardial infarction (AMI)

**Background:** Cardiovascular diseases (CVDs) are the leading cause of death globally. In 2019, an estimated 17.9 million people died from CVDs, accounting for 32% of all global deaths. Among these deaths, 85% were due to heart attacks and strokes.

Left ventricular ejection fraction (LVEF) recovery after myocardial infarction (MI) is an important prognostic indicator, and patients who do not recover LVEF after MI are at high risk of sudden cardiac arrest events and death.

Stem cell therapy holds promise for cardiovascular diseases, offering regenerative potential through cell differentiation. However, limited access exists for clinicians and patients. This study investigates the safety and efficacy of MiSaver (Myocardial Infarction Functional Saver), a prefabricated stem cell investigational product, in recent AMI patients. Findings contribute to advancing stem cell treatments, improving accessibility and patient outcomes.

**Methods:** Patients who were admitted for AMI within 7 days and had reduced LVEF (.45%) were eligible for the study. MiSaver were matched for blood group and administered in participants in cohorts of five, each receiving escalating dosages (0.5×10^7, 1.6×10^7, and 5.0×10^7 cells/kg, respectively). Patients were assessed for symptoms of graft-versus-host disease (GVHD) and treatment-related adverse events (AE). LVEF measured by echocardiographic on admission, at 6 months, and at 12 months after treatment. Patients functional activity status evalution ( using the New York Heart Association (NYHA) and Canadian Cardiovascular Society (CCS) classification systems.

**Results:** Out of the initially planned 15 participants, eleven were enrolled in the study. The trial was halted prematurely due to challenges associated with the COVID-19 pandemic and impractical transportation logistics. Patients received MiSaver infusions within 2-5 days post-AMI onset. During the 12-month follow-up period, no study-related adverse events or signs of graft-versus-host disease were reported. At 12 months post-treatment, both the low and middle dose groups, as well as participant 11, showed improved LVEF, accompanied by enhanced Canadian Cardiovascular Society (CCS) class grades compared to baseline.

**Conclusion:** The intravenous infusion of MiSaver stem cells in AMI patients demonstrated safety and tolerability for low and middle dosage groups. The study provides promising insights into the potential of stem cells therapy in improving left ventricular function following AMI. However, further research with larger cohorts and a controlled placebo is warranted to confirm these findings and address limitations encountered during this trial.

## Introduction

Cardiovascular diseases are the leading cause of death globally, resulting in an estimated 17.9 million deaths in 2019, with 85% of these deaths caused by myocardial infarction (MI) and stroke [1]. Despite advances in pharmacological and nonpharmacological treatments, MI remains one of the leading causes of death worldwide, with significant associated morbidity [2]. Consequently, numerous efforts have been made to develop alternative or novel treatments to improve cardiac functions and recovery.

Stem cells have emerged as a promising novel option for cardiac regeneration and re-vascularization, given their ability to migrate and modulate the immune system, secrete trophic factors to enhance angiogenesis, differentiate into mature functioning cells such as cardiomyocytes and vascular endothelial cells, reduce infarct volume, and improve cardiomyopathy outcomes [4-10].

MiSaver stem cell injections, derived from umbilical cord blood, are inherently advantageous as they are immuno-naive, immunologically tolerant, contain assorted types of non-hematologic stem cells such as endothelial progenitor stem cells, mesenchymal stem cells, multilineage progenitor cells, neural precursor stem cells, unrestricted somatic stem cells, and very small embryonic-like stem cells. Furthermore, they have been shown to be clinically safe through their hematological use in allogeneic hematopoietic stem cell transplantation for over two decades [11-17].

Several clinical trials have indicated the beneficial effects of stem cells on myocardial function in patients [18-20]. The degree of LVEF recovery after MI provides important prognostic information, with patients who do not experience any recovery at high risk of sudden cardiac arrest events and death [3]. This trial evaluated the safety of MiSaver and preliminary effectiveness for LVEF recovery in AMI patients.

## Methods

### Study Design and Overview

An open-label, dose-escalating clinical trial was conducted at Chung Shan Medical University Hospital to assess the safety and preliminary effectiveness of MiSaver stem cells in improving LVEF in patients with AMI. The study product consisted of ABO matched allogeneic umbilical cord blood stem cells (USC) prefabricated into MiSaver. Approval was obtained from the institutional review board of the clinical site, and regulatory authorities overseeing clinical trials and healthcare products.

### Patients

The eligibility criteria for participation in this study were carefully chosen to ensure the safety and relevance of the results. Patients who met the following criteria were eligible: aged between 20 and 80 years, diagnosed with AMI within 7 days, with elevated cardiac enzymes (CK-MB or troponin) greater than 2 times the high-end of normal value, presence of regional wall motion abnormality, and LVEF of ≤ 45% on clinical echocardiography.

It should be noted that patients were required to be hemodynamically stable, with no need for inotropic support, a systolic blood pressure lower than 80mm Hg for less than 1 hour, and a resting heart rate of >100 beats/min for less than 1 hour during the past 24-hour period. Additionally, patients were required to have a peripheral artery oxygen saturation of ≥97%.

Patients who were pregnant or breastfeeding, had a positive adventitious infection (such as HIV, hepatitis), required revascularization via coronary artery bypass surgery or anticipated further coronary revascularization procedures during the 6-month study period, had severe aortic or mitral valve narrowing, or had evidence of life-threatening arrhythmia on baseline electrocardiogram (ECG) were excluded. Patients who were short of breath and unable to receive PCI examination or treatment (including New York Heart Association, NYHA, Fc.IV), had a malignant tumor, hematopoietic dysplasia, or other severe organ disease, had less than 1 year of life expectancy, or had chronic kidney disease with CCr<20ml/min and were on renal dialysis were also excluded from the study.

Participants were enrolled in cohorts of five, each receiving escalating dosages (0.5×10^7, 1.6×10^7, and 5.0×10^7 cells/kg, respectively) of MiSaver stem cells via intravenous infusion 2-5 days post-AMI onset. Prior to stem cell therapy, all participants received standard treatment for AMI, including medication, percutaneous coronary intervention (PCI) with or without stent implantation.

### Stem Cells

USC were requested under the name of MiSaver from a GMP standard stem cell pharmaceutical company in Taiwan (HONYA Medical PTY LTD), based on blood type and the following specifications: (a) 200 million total nucleated cell count (TNC) in a 13ml solution, packaged in single-use 20ml clear vials with coated stopper seals and flip-off caps; (b) viability greater than 80%; (c) negative results for bacterial and fungal cultures; (d) negative nucleic acid testing (NAT) for infectious diseases including HIV, hepatitis C virus, hepatitis B virus (HBV), as well as negative antibody testing for syphilis; and (e) negative results for endotoxin testing.

### Administrations

MiSaver was thawed using a thermostat-controlled mini bath, drawn into a 20ml syringe, and diluted with normal saline and intravenous infused following standard operating procedures.

Patients were premedicated with intravenous antihistamines (such as diphenhydramine) and corticosteroids (such as hydrocortisone) 30–60 minutes before the stem cell infusion. Any unused solution was discarded, and the total volume of injected solution was adjusted to accommodate the total daily fluid volume administered.

### Safety Evaluation & Parameters

During hospitalization (from D0 to D3), and at scheduled follow-ups 1 month, 2 months, 3 months, 6 months, 9 months and 12 months after treatment, patients were closely monitored for safety. Throughout the treatment, heart rate, respiratory rate and oxygen saturation were continuously monitored. Prior to the infusion, as well as at 15-minute intervals for 1 hour post-infusion, and at 30-minute intervals for 2 hours post-infusion, physiological parameters including heart rate, blood pressure, temperature, respiratory rate, and oxygen saturation were reviewed for any AEs or changes to the patients’ physiological statuses.

To evaluate the response and reaction of the myocardium after cell therapy, serum levels of troponin and creatine kinase were tested daily from the day of treatment until a noticeable trend of decreasing level.

At follow-up visits, patients were assessed for symptoms of graft-versus-host disease (GVHD), infection, AEs, laboratory examinations, and ECG. Direct and indirect Coombs were tested at enrollment and 3 months post-treatment, while 24-hour Holter ECG and echocardiography were performed before the treatment, at 6 and 12 months after the treatment. Additionally, MRI scans were done before the treatment and 12 months after the treatment.

Procedural complications were defined as any ventricular arrhythmia, visible thrombus formation, distal embolization, or injury of the coronary artery associated with the stem cell treatment procedure.

### Statistical Analysis

Quantitative data are reported as median (25th percentile - 75th percentile), while qualitative data are presented as absolute frequencies and/or percentages. Fisher’s exact test was used to test differences between therapy groups for qualitative variables due to the small number of patients in each group. The non-normal distribution of the quantitative variables was identified, and we used the Wilcoxon signed-rank test and Wilcoxon rank-sum test to compare within-group differences and differences between therapy groups, respectively. The stepdown Bonferroni corrected p value was estimated for multiple comparisons. Results were considered statistically significant if the corresponding two-sided p-value was less than 0.05. Statistical analysis was conducted using SAS 9.4 (SAS Institute Inc., Cary, NC) software.

### Endpoints

The primary objective of this open-label phase 1 trial was to evaluate the safety profile of MiSaver stem cells by monitoring the incidence of study-related adverse events (AEs) and graft-versus-host disease (GVHD) and change in LVEF using echocardiography over a 12-month follow-up period. The secondary endpoints were to evaluate the functional activity status at admission and at 1, 2, 3, 6, 9 and 12 months post-treatment using the New York Heart Association (NYHA) and Canadian Cardiovascular Society (CCS) classification systems.

## Results

The study was initially to accrue 15 patients, but was closed early after 11 patients due to COVID pandemic and impractical transportation logistics. One participant was missing two post infusion physiological parameter figures because he was sent for examinations. One participant was missing some of the month 6 follow up blood results because insufficient blood specimen was collected at the follow up clinic.

### Baseline Characteristics of The Participants

Eleven male adult patients (median 56.2, range: 41-70 years) were enrolled in the MiSaver study between January 2021 and March 2022. Most participants had risk factors for MI, including hypertension, hyperlipidemia, diabetes mellitus, and a history of smoking (Figure 1).

**Figure 1.**
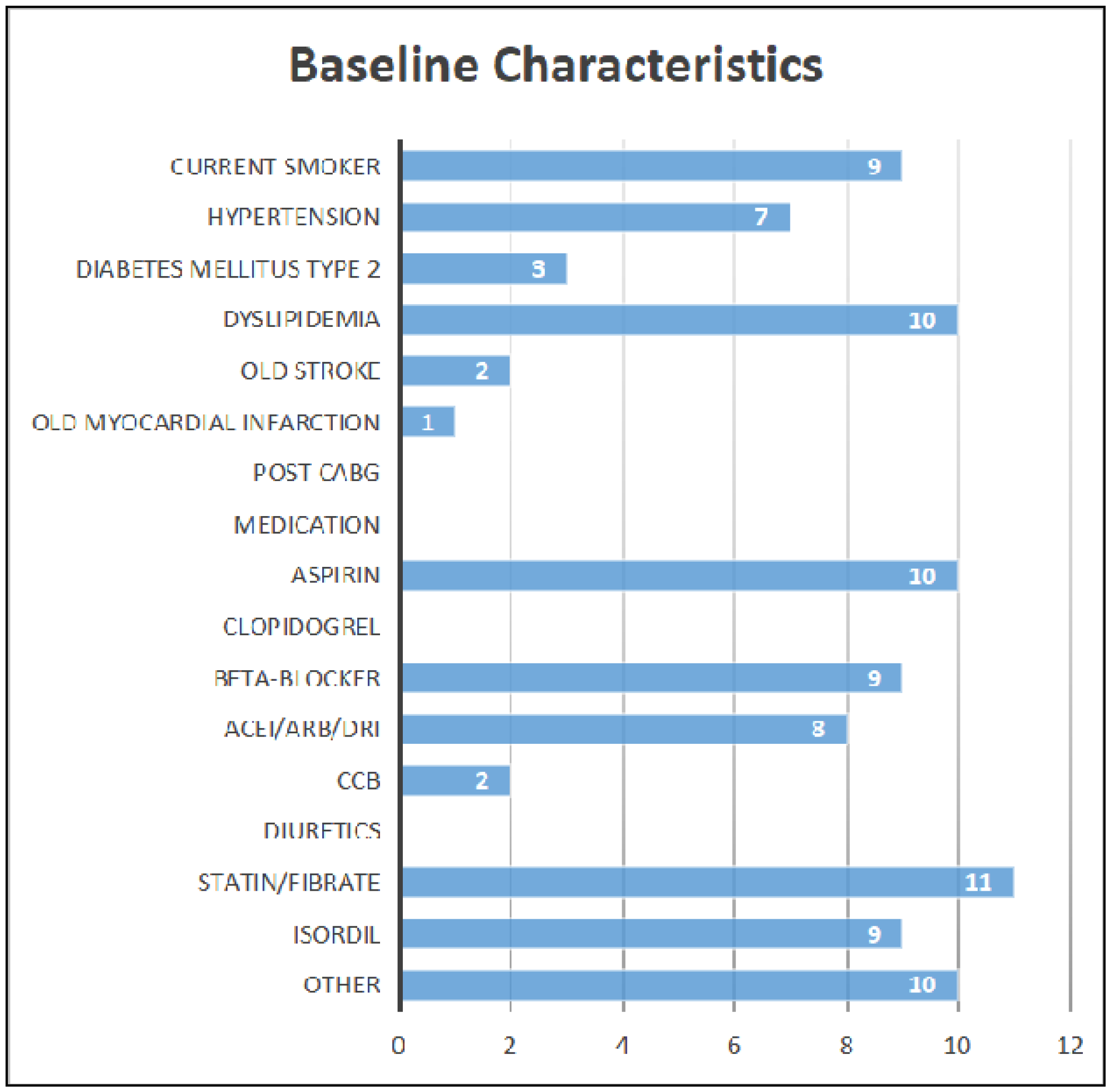
Baseline characteristics of participants.

### MiSaver Infusion

Before the infusion, clinical laboratory tests and echocardiography were conducted and reviewed. Baseline MRI and 24-hour Holter EKG were obtained prior to the infusion. Participants were administered with 0.5, 1.6 or 5.0 x10^7/kg cell dose of ABO/Rh matched MiSaver stem cells intravenously, within 2-5 days post MI as indicated in Table 1.

**Table 1.** Baseline characteristics of participants age, weight and MiSaver infusion time and dosages.

| Participant | Age | Infusion,<br>days post MI | Body weight (kg) | MiSaver Cell Dose x10<sup>7</sup>/kg |
| --- | --- | --- | --- | --- |
| 1 | 66~70 | 4 | 70.4 | 0.5 |
| 2 | 41~45 | 2 | 79.4 | 0.5 |
| 3 | 56~60 | 3 | 62 | 0.5 |
| 4 | 51~55 | 3 | 56.8 | 0.5 |
| 5 | 56~60 | 2 | 76 | 0.5 |
| 6 | 66~70 | 4 | 59.6 | 1.6 |
| 7 | 41~45 | 2 | 97 | 1.6 |
| 8 | 66~70 | 5 | 66.55 | 1.6 |
| 9 | 41~45 | 3 | 75 | 1.6 |
| 10 | 41~45 | 3 | 100 | 1.6 |
| 11 | 61~65 | 3 | 78 | 5.0 |
| Median(Range) | 60(41~70) | 3 (2~5) | 75 (56.8~100) | 1.6 (0.5~5.0) |

**Table 2.**
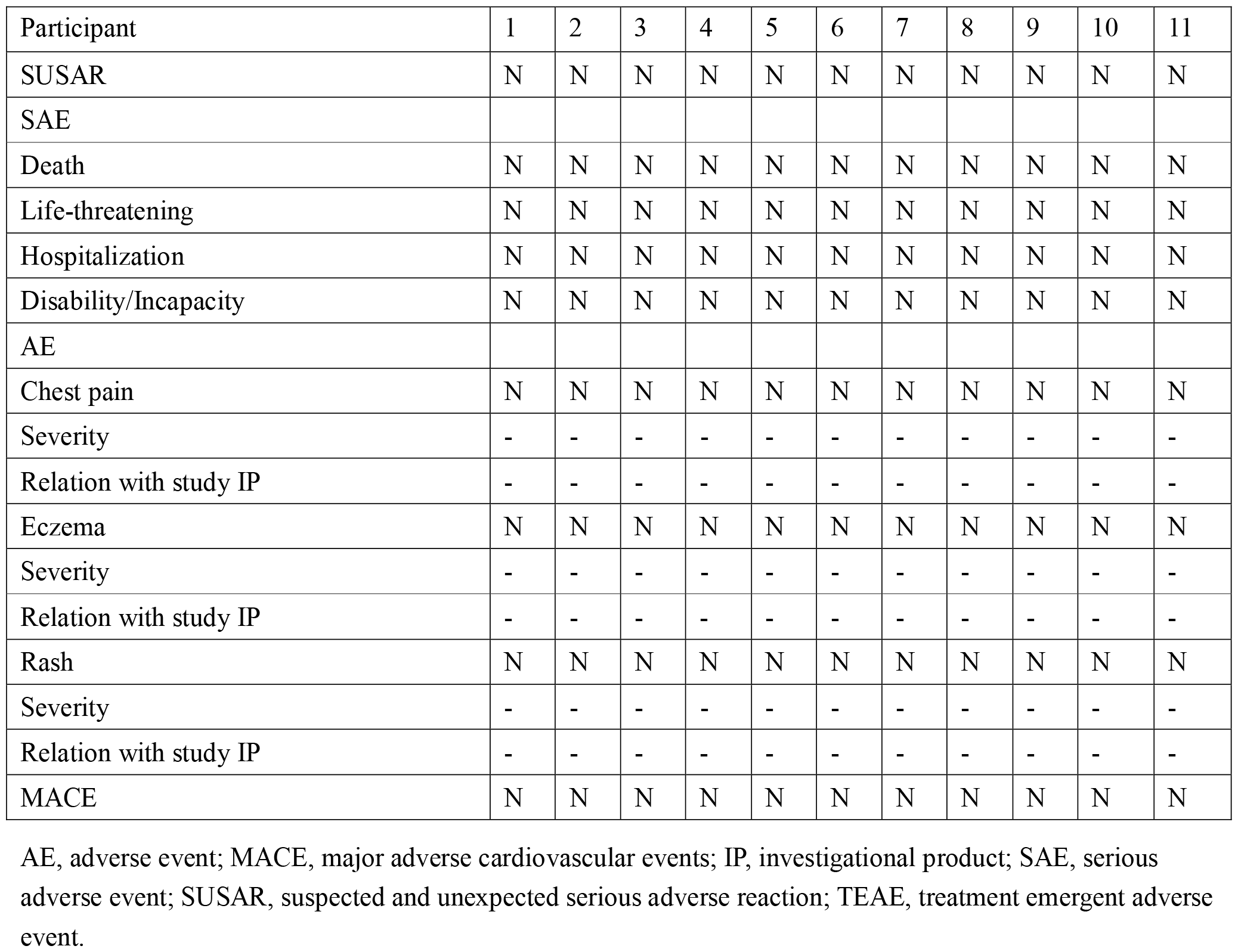
Adjudication of treatment related adverse events among study subjects.

### Safety Assessment

The primary aim of this open-label phase 1 trial was to evaluate the safety profile of MiSaver stem cells by monitoring the incidence of study-related adverse events (AEs) and graft-versus-host disease (GVHD) over a 12-month follow-up period. Notably, there were no study-related AEs reported.

Participant 9, at 12 months post-treatment, visited the emergency department due to fever and subsequently tested positive for COVID-19. After receiving symptom management, the participant’s condition stabilized, and returned home on the same day. This incident was concluded unrelated to the investigational drug.

Participant 10, approximately 2 and a half months post-treatment, suspected a hospital-acquired COVID-19 infection while caring for his mother in the hospital. He underwent testing and isolation at the epidemiology outpatient clinic. This incident was deemed unrelated to the investigational drug. The participant also delayed his return visit by a few days and completed the 2-month follow-up. Additionally, the participant reported experiencing diarrhea a few days prior to the 12 month follow up visit, which resolved spontaneously without medication by the time of the visit. This event was considered unrelated to the investigational drug.

On day 4, Participant 11, who received a high dose of 5.0 x 10^7/kg cells, showed a slight elevation in Troponin levels, rising from 11052 to 13037, while CK-MB levels were recorded as

7.2 and 7.4. By day 5, both Troponin and CK-MB levels decreased to 8723 and 6.3, respectively. This event was duly documented and monitored as part of the safety evaluation process.

### Echocardiogram Analysis

Both the low (L) and middle (M) dose treatment groups improved in LVEF compared to baseline (L: 42 [38.3-42.3], M: 40.65 [33.95-41.95]) at both 6 months (L: 49.6% [48.3-52.1], M: 53 [47.5-61.8]) and 12 months (L: 53% [46.1-68.6], M: 54.7 [48.15-55.9]). Importantly, no statistically significant differences were noted between the two groups at baseline, 6 months, or 12 months (p=0.4567, 0.3886, or 0.5871, respectively) (Figure 2). Furthermore, the combined treatment groups (L and M dose groups) exhibited a statistically significant increase in LVEF from baseline (41.7%) to 6 months (50.4%) and a further improvement, although not statistically significant, to 53.4% at 12 months (refer to Figure 3).

**Figure 2.**
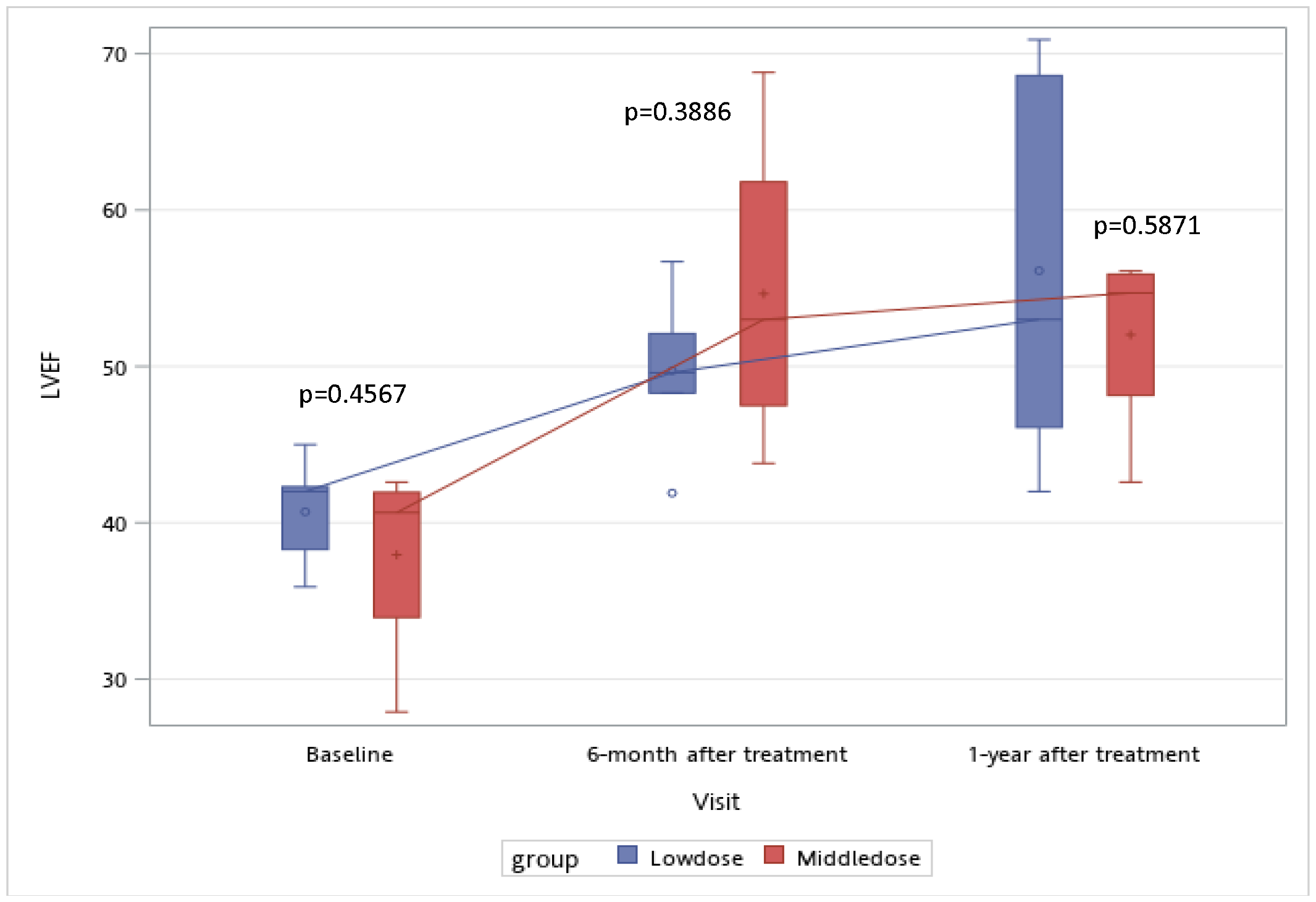
LVEF values for low (L) and middle (M) dosage groups. Figure 2, shows the mean LVEF values at baseline, 6 months, and 12 months for low (L) and middle (M) dosage groups. Both groups experienced an increase in LVEF from baseline (L: 42 (38.3-42.3), M: 40.65 (33.95-41.95)) to 6 months (L: 49.6% (48.3-52.1), M: 53 (47.5-61.8)) and to 12 months (L: 53% (46.1-68.6), M: 54.7 (48.15-55.9)). There was no statistical difference between the two groups at baseline, 6 months, or 12 months (p = 0.4567, 0.3886, or 0.5871, respectively).

**Figure 3.**
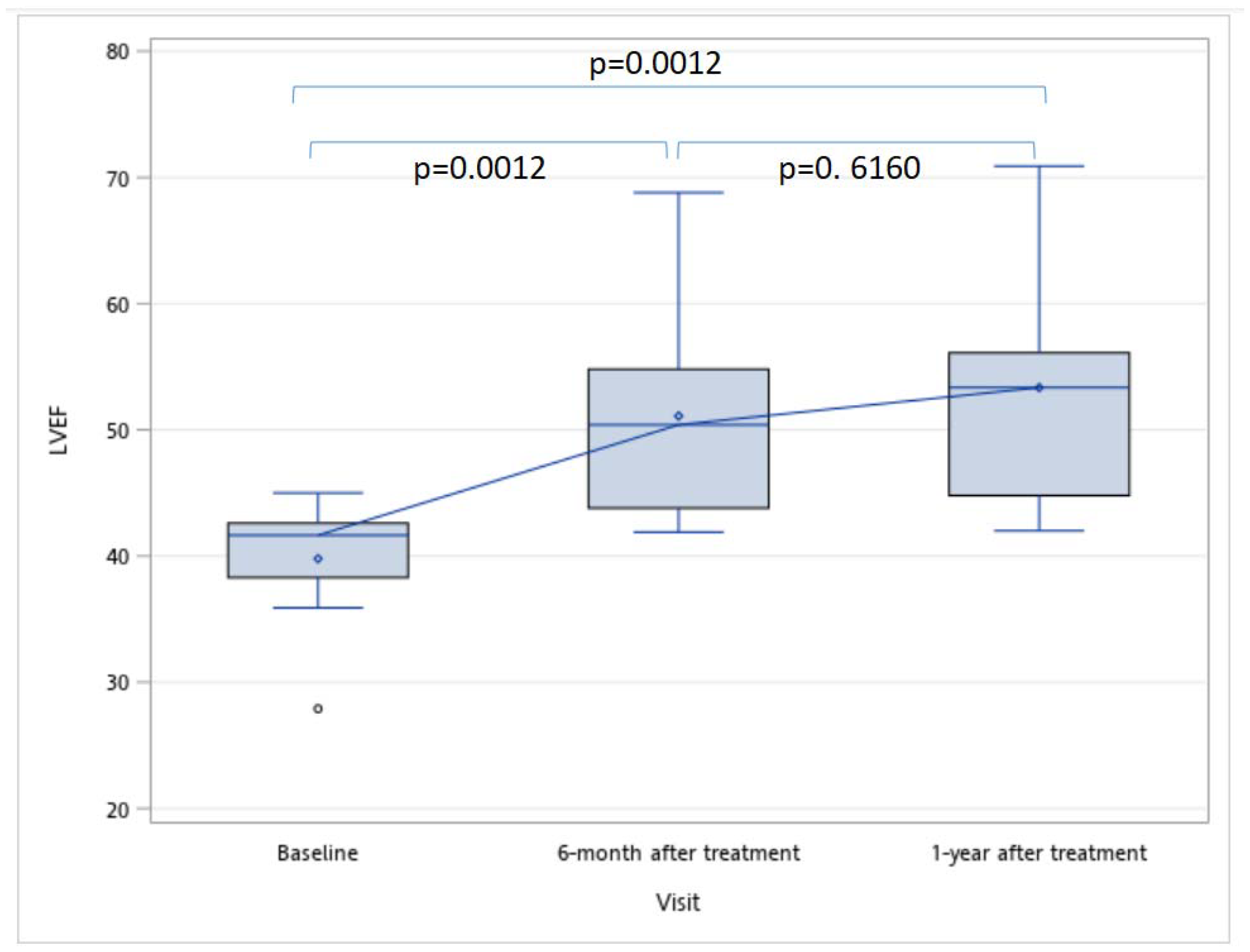
LVEF of treatment group (L+M) at base line, 6 months and 12 months.

### Holter EKG Monitoring

Participants underwent Holter EKG monitoring for a median duration of 22 hours and 53 minutes, ranging from 3 hours 1 minute to 23 hours 59 minutes. The average median heart rate across all participants was 78 beats per minute, with a maximum heart rate of 135 beats per minute recorded in Participant 7 at month 12. Conversely, Participant 2 exhibited a minimum heart rate of 24 beats per minute at month 6. Additionally, Participant 1 recorded a maximum of 9.6 ventricular beats at month 6, although many of these beats were identified as artifacts (see Table 3). Importantly, there were no symptomatic complaints associated with the observed tachycardia, bradycardia, or arrhythmic signs.

**Table 3.**
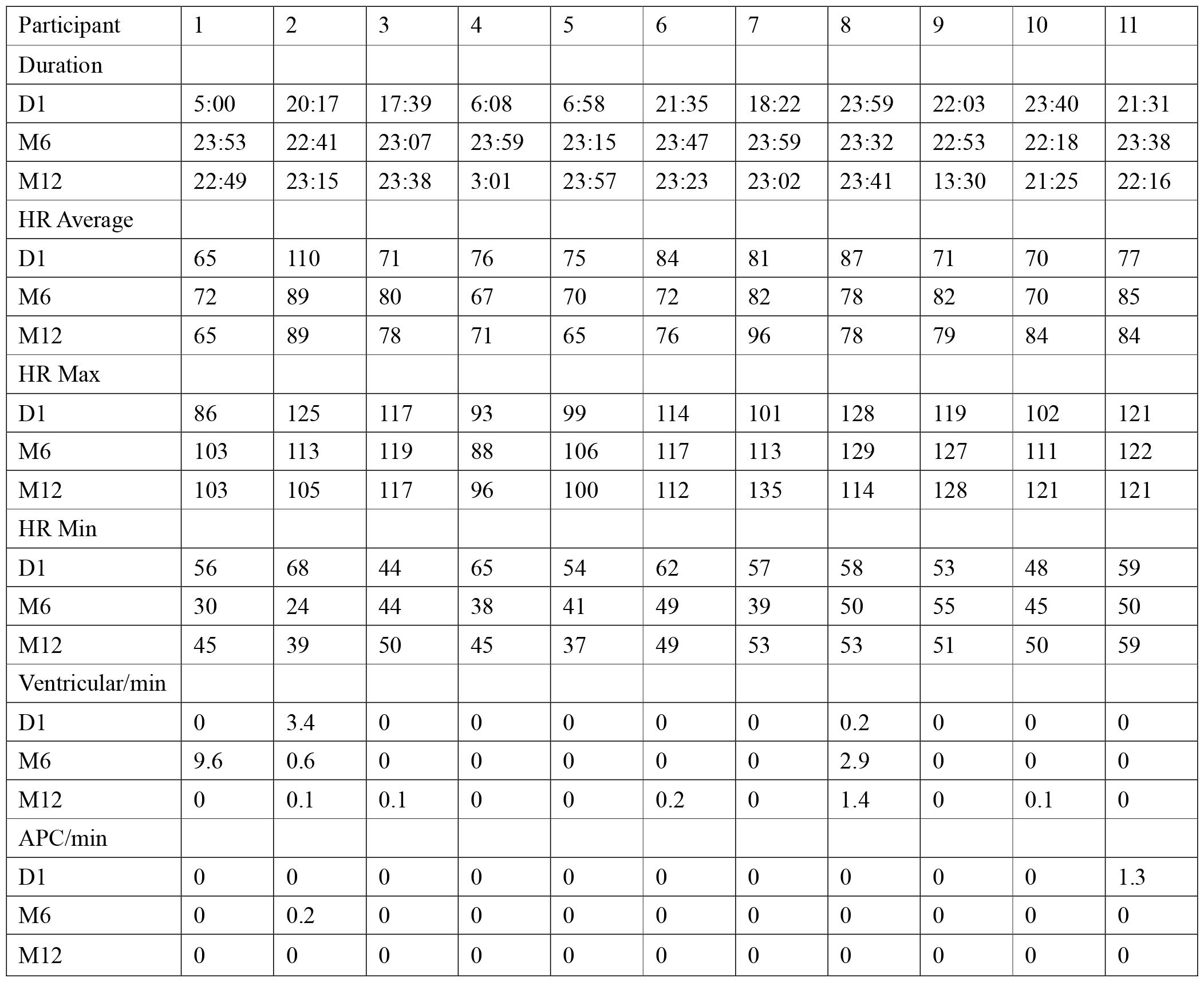
Holter EKG Result.

### Immune Assay

All participants underwent comprehensive immune assays. Participant 1’s Coombs tests showed a seroconversion from positive to negative, while Participant 11 experienced a conversion from negative to positive in the Indirect Coombs test. Additionally, Participant 11 exhibited a positive conversion in the Mixed Lymphocyte Reaction test. Apart from inadequate specimen for laboratory testing from Participant 2 in the Panel Reactive Antibody Screening, no other instances of positive conversions were detected for either the Mixed Lymphocyte Reaction or Panel Reactive Antibody Screening (see Table 4).

**Table 4.** Immunological tests during the study period.

| Table 4. Immunological tests during the study period |  |  |  |  |  |  |  |  |  |  |  |
| --- | --- | --- | --- | --- | --- | --- | --- | --- | --- | --- | --- |
| Participant | 1 | 2 | 3 | 4 | 5 | 6 | 7 | 8 | 9 | 10 | 11 |
| Direct Coombs |  |  |  |  |  |  |  |  |  |  |  |
| D1 | Pos | Neg | Neg | Neg | Neg | Neg | Neg | Neg | Neg | Neg | Neg |
| M3 | Neg | Neg | Neg | Neg | Neg | Neg | Neg | Neg | Neg | Neg | Neg |
| Indirect Coombs |  |  |  |  |  |  |  |  |  |  |  |
| D1 | Neg | Neg | Neg | Neg | Neg | Neg | Neg | Neg | Neg | Neg | Neg |
| M3 | Neg | Neg | Neg | Neg | Neg | Neg | Neg | Neg | Neg | Neg | Pos |
| Mixed lymphocyte reaction |  |  |  |  |  |  |  |  |  |  |  |
| D1 | Pos | Pos | Neg | Neg | Pos | Neg | Neg | Neg | Neg | Neg | Neg |
| M3 | Pos | I | Neg | Neg | Pos | Neg | Neg | Neg | Neg | Neg | Pos |
| Panel reactive antibody screening |  |  |  |  |  |  |  |  |  |  |  |
| D1 | Pos | I | Neg | Neg | Neg | Neg | Neg | Neg | Neg | Neg | Neg |
| M3 | I | I | Neg | Neg | Neg | Neg | Neg | Neg | Neg | Neg | Neg |
| Pos: positive; Neg: Negative; I : inadequate specimen |  |  |  |  |  |  |  |  |  |  |  |

### NYHA and CCS Functional Classification Outcomes

All participants exhibited improvements in the CCS functional classification activities. Initially, seven participants were classified as CCS II, while four participants were classified as CCS I. Over the 12-month period, seven participants demonstrated a one-functional-class improvement, and four participants displayed a two-functional-class improvement.

In terms of the NYHA classification, ten participants were categorized as NYHA II, and one participant was categorized as NYHA I. Throughout the 12-month period, seven participants experienced a one-functional-class improvement, while two participants experienced a two-functional-class improvement.

## Discussion

This clinical trial aimed to evaluate the safety and preliminary effectiveness of MiSaver, an investigational stem cell product designed to provide clinicians with a convenient method of accessing stem cells without the need for extensive laboratory facilities.

Participants were divided into cohorts of five, each receiving escalating dosages (0.5×10^7, 1.6×10^7, and 5.0×10^7 cells/kg, respectively), with infusions administered 2-5 days post-AMI onset. Of the planned 15 participants, 11 were included in the primary analyses, and the trial was terminated prematurely due to challenges associated with the COVID-19 pandemic and logistical issues.

Over the 12-month follow-up period, no study-related adverse events were reported, no signs or symptoms of GVHD were observed, and there were no instances of AMI recurrence or re-hospitalization among the participants. These findings suggest that MiSaver is safe and well-tolerated in AMI patients.

The combined treatment groups (L and M dose groups) demonstrated a total increase of 11.7% in left ventricular ejection fraction (LVEF) over the 12-month period. Although the middle dose group exhibited a 3.4% higher improvement in LVEF compared to the low dose group at 6 months, this difference did not reach statistical significance. Therefore, further investigation in larger clinical trials with controlled parameters is warranted to determine the significance of these findings.

All participants in the study showed improvement in the CCS functional classification scale. However, to accurately assess the significance of this improvement, further investigation with a controlled placebo group is recommended.

Given the safety profile, preliminary efficacy results, and ease of administration of MiSaver stem cells, larger-scale studies are necessary to confirm these findings.

## Conclusion

Based on the findings of this phase I trial, it can be concluded that intravenous infusion of MiSaver stem cells in patients with AMI is safe and well-tolerated. These findings provide encouraging evidence for the continued development and investigation of MiSaver stem cells as a potential therapy for AMI. However, further research is needed to explore any potential differences and their significance in larger-scale clinical trials.

### Limitations

This study has several limitations that may impact the interpretation of the results. Firstly, only male patients were enrolled in the study, despite both sexes being eligible to participate, potentially limiting the generalizability of the findings. Secondly, due to the COVID-19 pandemic during the trial period, only 11 out of the planned 15 patients were enrolled, and the study was prematurely closed. Thirdly, participants were subject to quarantine during the study period, which may have influenced the exact follow-up time or results. Future studies should aim to address these limitations to improve the generalizability and validity of the findings.

### Contributorship

K.U.: conception and design, provision of study material or patients, data analysis and interpretation, manuscript writing, final approval of manuscript; C.T., C.S. and Y.C. : provision of study material or patients; J.L.: conception and design, manuscript writing, collection and/or assembly of data, data analysis and interpretation.

### Ethical Approval and Compliance with the Declaration of Helsinki

This clinical trial was conducted in accordance with the ethical principles outlined in the Declaration of Helsinki. The research protocol received approval from the Institutional Review Board of Chung Shan Medical University Hospital, reference numbers CS19037. Informed consent was obtained from all participants (or their legal guardians), ensuring the protection of their rights, welfare, and privacy.

## Data Availability

All data produced in the present study are available upon reasonable request to the authors

## Acknowledgments and Special Thanks

This research was funded by Chung Shan University Hospital (Research Project: CSH-2018-C-005). Dr. J. Chen, head of Pojen OG Hospital, for facilitating and enabling the collection of stem cells. JY Huang for data analysis and interpretation.

## Funding, Grant/Award Info

This clinical trial is co-funded by Chung Shan University Hospital (Research Project: CSH-2018-C-005) and HONYA Medical.

## Data Sharing Statement

All data produced in the present study are available upon reasonable request to the authors.

## References

1. World Health Organization, statistics update 2019.

2. EJ Benjamin, et al. American Heart Association Council on Epidemiology and Prevention Statistics Committee and Stroke Statistics Subcommittee. Heart disease and stroke statistics-2019 update: a report from the American Heart Association. Circulation. 2019;139:e56–e528.

3. Derek S. Chew, et al. The degree of LVEF recovery after a first MI provides important prognostic information. Patients with no recovery in LVEF after MI are at high risk of sudden cardiac arrest events and death. J Am Coll Cardiol EP 2018;4:672–82.

4. C. van de Ven, D. Collins, M. B. Bradley, E. Morris, and M. S. Cairo, “The potential of umbilical cord blood multipotent stem cells for nonhematopoietic tissue and cell regeneration,” Experimental Hematology, vol. 35, no. 12, pp. 1753–1765, 2007.

5. G. Kogler, S. Sensken, J. A. Airey et al., “A new human somatic stem cell from placental cord blood with intrinsic pluripotent differentiation potential,” Journal of Experimental Medicine, vol. 200, no. 2, pp. 123–135, 2004.

6. Psaltis PJ, Paton S, See F, Arthur A, Martin S, Itescu S, Worthley SG, Gronthos S, Zannettino AC. Enrichment for STRO-1 expression enhances the cardiovascular paracrine activity of human bone marrow-derived mesenchymal cell populations. J Cell Physiol. 2010; 223:530–540. doi:10.1002/jcp.22081.

7. Kim YJ, Broxmeyer HE. Immune regulatory cells in umbilical cord blood and their potential roles in transplantation tolerance. Crit Rev Oncol Hematol 2011; 79:112–126.

8. Gnecchi M, Zhang Z, Ni A, Dzau VJ. Paracrine mechanisms in adult stem cell signaling and therapy. Circ Res. 2008; 103:1204–1219. doi:

9. Huang NF, Li S. Mesenchymal stem cells for vascular regeneration. Regen Med. 2008; 3:877– 892. doi:10.2217/17460751.3.6.877.

10. Psaltis PJ, Carbone A, Nelson AJ, Lau DH, Jantzen T, Manavis J, Williams K, Itescu S, Sanders P, Gronthos S, Zannettino AC, Worthley SG. Reparative effects of allogeneic mesenchymal precursor cells delivered transendocardially in experimental nonischemic cardiomyopathy. JACC Cardiovasc Interv.

11. R. Vyas, D. Dudhat, N. Sudhalkar, V. Garg, J. Thadani, A. Marathe, et al. Clinical safety in using unmatched allogeneic umbilical cord blood mononuclear cells transplantation in non-haematopoietic degenerative conditions. Journal of Stem Cells, 9 (4) (2014), pp. 219–224

12. Yuan C, Yu G, Yang T. Enhanced therapeutic effects on acute myocardial infarction with multiple intravenous transplantation of human cord blood mononuclear cells Int J Cardiol. 2013; 168:2767–2773.

13. DT Laskowitz et al. Allogeneic Umbilical Cord Blood Infusion for Adults with Ischemic Stroke: Clinical Outcomes from a Phase I Safety Study. Stem Cell Translational Medicine, 2018;7:521–529.

14. Matsumoto and Mugishima. Non-Hematopoietic Stem Cells in Umbilical Cord Blood. International Journal of Stem Cells 2009;2:83–89.

15. Hows JM, Marsh JC, Bradley BA. Human cord blood: A source of transplantable stem cells? Bone Marrow Transplant. 1992;9(Suppl 1)105–108.

16. Munoz J, Shah N, Rezvani K. Concise review: Umbilical cord blood transplantation: Past, present, and future Stem Cells Translational Medicine. 2014; 3:1435–1443.

17. JE Wagner, Jr. Et. al. One-Unit versus Two-Unit Cord-Blood Transplantation for Hematologic Cancers. N Engl J Med. 2014 Oct 30; 371(18):1685–1694.

18. C Yuan,G Yu, T Yang. Enhanced therapeutic effects on acute myocardial infarction with multiple intravenous transplantation of human cord blood mononuclear cells. Int J Cardiol. 2013;168:2767–2773.

19. Meluzín J, Mayer J, Groch L. Autologous transplantation of mononuclear bone marrow cells in patients with acute myocardial infarction: The effect of the dose of transplanted cells on myocardial function Am Heart J. 2006;152:975. e9–975.e15.

20. Yuan C, Yu G, Yang T. Enhanced therapeutic effects on acute myocardial infarction with multiple intravenous transplantation of human cord blood mononuclear cells Int J Cardiol. 2013; 168:2767–2773.

21. Ghiroldi A, et al. Cell-based therapies for cardiac regeneration: a comprehensive review of past and ongoing strategies. Int J Mol Sci 2018;19:3194.

22. AT Ulus, et al. Intramyocardial Transplantation of Umbilical Cord Mesenchymal Stromal Cells in Chronic Ischemic Cardiomyopathy: A Controlled, Randomized Clinical Trial (HUC-HEART Trial). International Journal of Stem Cells Published online August 31, 2020.

23. MG Iachininoto, et al. In vitro cardiomyocyte differentiation of umbilical cord blood cells: crucial role for c-kit(+) cells. Cytotherapy. 2015 Nov;17(11):1627–37.

24. Rasha A.M. Khattab, et al. In vitro transdifffferentiation of umbilical cord stem cells into cardiac myocytes: Role of growth factors. The Egyptian Journal of Critical Care Medicine (2013) 1, 43–50.

25. N Nishiyama, et al. The Significant Cardiomyogenic Potential of Human Umbilical Cord Blood-Derived Mesenchymal Stem Cells In Vitro. STEM CELLS 2007;25:2017–2024.

